# Intubation and Inhospital mortality in trauma patients with Glasgow Coma Scale Score eight or less. A multicenter cohort study

**DOI:** 10.1101/2022.03.24.22272861

**Authors:** Kapil Dev Soni, Varun Bansal, Monty Khajanchi, Deepa Kizhakke Veetil, Anderson Geoffrey, Nakul Rayker, Bhakti Sarang, Siddharth David, Martin Gerdin Wärnberg, Nobhojit Roy

**Affiliations:** Critical and Intensive Care, JPN Apex Trauma Centre, AIIMS, New Delhi, India; Seth. G. S. Medical College & K.E.M. Hospital, Parel, Mumbai, India; Department of Surgery, Manipal hospital,New Delhi, India; Division of Trauma, Burn, Surgical Critical Care and Emergency General Surgery, Brigham & Women’s Hospital, Boston, USA; Division of Trauma and Emergency Surgery, Brigham and Women’s Hospital, Boston, Program in Global Surgery and Social Change, Harvard Medical School, Boston, USA; Trauma Research Group, WHO Collaborating Centre for Research in Surgical Care Delivery in LMICs, Mumbai, India; Doctors For You, Mumbai, India; Health Systems and Policy, Department of Global Public Health, Karolinska Institutet, Stockholm, Sweden; Department of Global Public Health, Karolinska Institutet, Stockholm, Sweden, Function Perioperative Medicine and Intensive Care, Karolinska University Hospital, Solna, Sweden; Trauma Research Group, WHO Collaborating Centre for Research in Surgical Care Delivery in LMICs, Mumbai, India, Dept of Global Public Health, Karolinska Institutet, Stockholm, Sweden, School of Public Health and Preventive Medicine, Monash University, Melbourne, Vic, Australia

**Keywords:** Trauma, Glasgow Coma Score, Injury, Intubation, Inhospital mortality

## Abstract

**Background:** Most trauma societies recommend intubation of trauma patients with Glasgow coma scale (GCS) ≤ 8, without robust evidence supporting it.

**Methods:** We examined the association between intubation and inhopital 30 day mortality in trauma patients arriving with GCS ≤ 8. The data were obtained using the Towards Improved Trauma Care Outcomes (TITCO) registry in India cohort. We compared the outcomes of patients with GCS ≤ 8 who were intubated within one hour after arrival with those who were intubated later or not at all, using multiple analytical approaches to evaluate the consistency of the findings. We also examined the association in multiple subgroups to identify any variability of the effect.

**Results:** Of 3476 patients who arrived with a GCS ≤ 8, 1671 (48.1%) were intubated within an hour and 1805 (51.9%) were intubated later or not intubated at all. Overall, 1957 (56.3%) patients died in whole cohort. A total of 947 (56.7%) patients died in intubation group and 1010 (56%) died in non intubation group. In the main analysis, there was no significant association between intubation within an hour and mortality(OR=1.18,[CI,0.76-1.84], p value = 0.467). This result was consistent across multiple sensitivity analysis.

**Conclusion:** In this observational study of trauma patients with GCS ≤ 8, who present to tertiary care hospitals, intubation within one hour after arrival was not associated with increased or decreased risk of inhospital mortality compared to intubation after one hour or no intubation. Further studies are needed to precisely evaluate the benefit of intubation and thus supporting the recommendations.

## Introduction

Airway management takes precedence in the stabilization of a patient with trauma in accordance with the ABCDE recommendation of the Advanced Trauma Life Support (ATLS) guidelines, based on the treat-first-that-kills-first principle. The American College of Surgeons (ACS) ATLS guidelines and the Eastern Association for the Surgery of Trauma (EAST) practice management guidelines (PMGs) recommend intubation of patients with a Glasgow Coma Scale (GCS) score ≤ 8, (1,2). However, the literature supporting this recommendation is insufficient, without any direct evidence of an outcome benefit to intubating all patients with GCS scores ≤ 8 arriving to hospital after acute injury. Albeit patient patients who arrive with GSC ≤ 8 for instance severe facial trauma, airway bleeding, traumatic cardiac arrest, injuries that warrant immediate surgery mandates immediate intubation but in other cases the need is uncertain. To date, there have been no robust clinical trials that have shown benefit of intubation in the specific patient cohort of GCS ≤ 8,. The data that come from earlier observational studies also failed to support the recommendation.(3,4)

Securing the airway following trauma is presumed to be a relatively safe procedure and it is often performed early in group of patients arriving with GCS ≤ 8 to mitigate the risk of hypoxia, ventilatory failure and aspiration. Although intubation preserves oxygenation and ventilation, it puts critically injured patients at further risk of hemodynamic imbalance and pulmonary compromise, as well as long term morbidity because of ventilator-associated complications.(5,6) Furthermore, the procedure is technically challenging in the acute trauma settings and if not performed correctly may lead to worse outcomes (7). Indeed, previous studies in which intubation was performed by the emergency technician pre hospital settings, lead to poor survival (8).

Adherence to guidelines regarding intubation is largely influenced by provider preference, resulting in significant interpersonal and center-to-center variation in airway management practices (9–11).These variation in practices of intubation are further amplified in resource limited settings, leading to reduced compliance and uncertainty regarding efficacy of the procedure. The timing of intubation may be a critical factor determining the overall benefits, as delay may be associated with reduced effectiveness Therefore in this study, we examined the association between intubation of patients with GCS≤ 8 and in hospital mortality..

## Materials and Methods

### Design and Settings

This study is an analysis of patients arriving after trauma with a GCS score ≤ 8 enrolled in the ‘Towards improving trauma care outcome (TITCO) cohort. (https://www.titco.org) This is a prospective cohort of trauma patients admitted to four urban university-based hospitals in India, from 1^st^ September 2013 to 31^st^ October 2015.The participating hospitals across three Indian states were: the Jai Prakash Narayan Apex Trauma Centre of the All-India Institute of Medical Sciences (AIIMS), New Delhi; King Edward Memorial Hospital (KEMH), Mumbai; Lokmanya Tilak Municipal General Hospital (LTMGH), Mumbai; and Seth Sukhlal Karnani Memorial Hospital (SSKM), Kolkata. These government hospitals are referral centers for tertiary trauma care for neighboring peripheral hospitals and states and serve by providing services free or for minimal charges. The Apex trauma center is a stand-alone center, whereas the others provide trauma services as a part of a general hospital. Two of the centers had a dedicated trauma bay, managed by general surgeons and other specialists, while other two managed all surgical emergencies (trauma and non-trauma) in the emergency surgery service (ESS) area. There is no organized pre- hospital care or any pre-hospital notification received in these hospitals (12). After referral from the receiving triage area of the hospital to the trauma bay/ESS, patients are stabilized by the intern physician and general surgery resident doctor. If deemed necessary, intubation is performed by an anaesthetist, intensivist, or general surgeon. Intensive care unit (ICU) care is variable across the centres and often limited by the availability of beds.

Data for the registry were collected by project officers located at the four centers in eight-hour shifts on a rotational basis. The data of patients admitted outside their duty shifts was recorded from the case records. The methodology has been explained in detail previously (13).

### Patients

For our analysis we included all patients, both adults and pediatric enrolled in the TITCO cohort presenting with GCS ≤ 8 and with complete data for the primary outcome,. The cohort includes all consecutive patients who presented after major trauma and who were admitted to the hospitals. Patients who were intubated or airway secured before arrival were considered intubated within hour group. It excludes patients who were dead on arrival or presented with low-energy injuries, such as single bone fractures, or who were not admitted in the participating centers.

### Variables

We analyzed the following variables : age, sex, mechanisms of injury (road traffic injuries, falls, railway injuries, physical assaults, burns, and others) (blunt VS penetrating mechanism), injury severity defined by the Injury Severity score (ISS) (14),transfer status (whether the patient was transferred from another facility to a study hospital or arrived directly), and mode of transfer. Vital parameters on arrival, including heart rate (HR), respiratory rate (RR) systolic blood pressure (SBP), and oxygen saturation (SpO2), as well as GCS scores, were also analyzed as quantitative variables. The length of ICU stay and length of ventilation if applicable, were documented as quantitative variables.

### Outcomes

The primary outcome of this study was to compare in-hospital mortality(30 day mortality) in patients intubated within one hour of hospital arrival and those not intubated within one hour or intubated later. The secondary outcomes were to compare the duration of hospitalization, ventilation, and ICU stay if applicable, as surrogates of morbidity.

### Statistical analysis

Descriptive statistics are presented for the overall cohort of patients. We calculated the bivariate frequencies to evaluate the associations among the baseline demographic variables and intubation status. Categorical variables are summarized using proportions. Quantitative variables are represented as by their median values and interquartile range.

The effect of intubation status on in-hospital mortality was assessed in an unadjusted analysis and after adjusting for age, SBP, GCS scores, and ISS, all of which were included as quantitative variables in a multivariable logistic regression model. The selection of variables were based on a priori prognostic considerations. Variables in the model were removed if evidence of collinearity was present.For logistic regression analysis only variables with complete observations were included to limit bias due to missing observations. A p-value ≤ 0.05 was considered significant with 95% CI.

In addition, to account for non-randomized intervention (Intubation) administration, propensity score methods were used to reduce confounding. The individual propensities for receiving of intubation were estimated using regression model that included the same covariates as describe above. Nearest neighbor method was applied to create a matched control sample and propensity matched analysis was performed

We also conducted a subgroup analysis to assess the outcomes of interest in patients with GCS scores of 6-8 and with scores 3-5, as these groups differ fundamentally in their response to treatment and likelihood of survival following trauma (15,16). An additional subgroup analysis was performed based on presence of isolated traumatic brain injury (TBI) and without TBI.To limit potential immortal bias arising from patients who were transferred from other hospitals, we also performed sensisitivity analyses by comparing mortality between the study groups by restricting analysis to patients who were admitted directly to these centers.

Lastly, the factors increasing the likelihood of the clinical decision to intubate in our cohort were evaluated, the included factors being age, SBP, SpO2, GCS, ISS, RR, and HR, and the fitness of the model being judged by the Area Under the Curve (AUC). Statistical analysis was performed using R software.

This study is reported in accordance with the Strengthening the Reporting of Observational Studies in Epidemiology (STROBE) guidelines. [Von Elm 2007]

## Results

### Characteristics of the Cohort

Among annonymoized dataset of 16047 patients in the TITCO registry, 3476 patients with GCS scores ≤ 8 met the inclusion criteria (Figure 1) and were included in the analysis.

**Figure 1:**
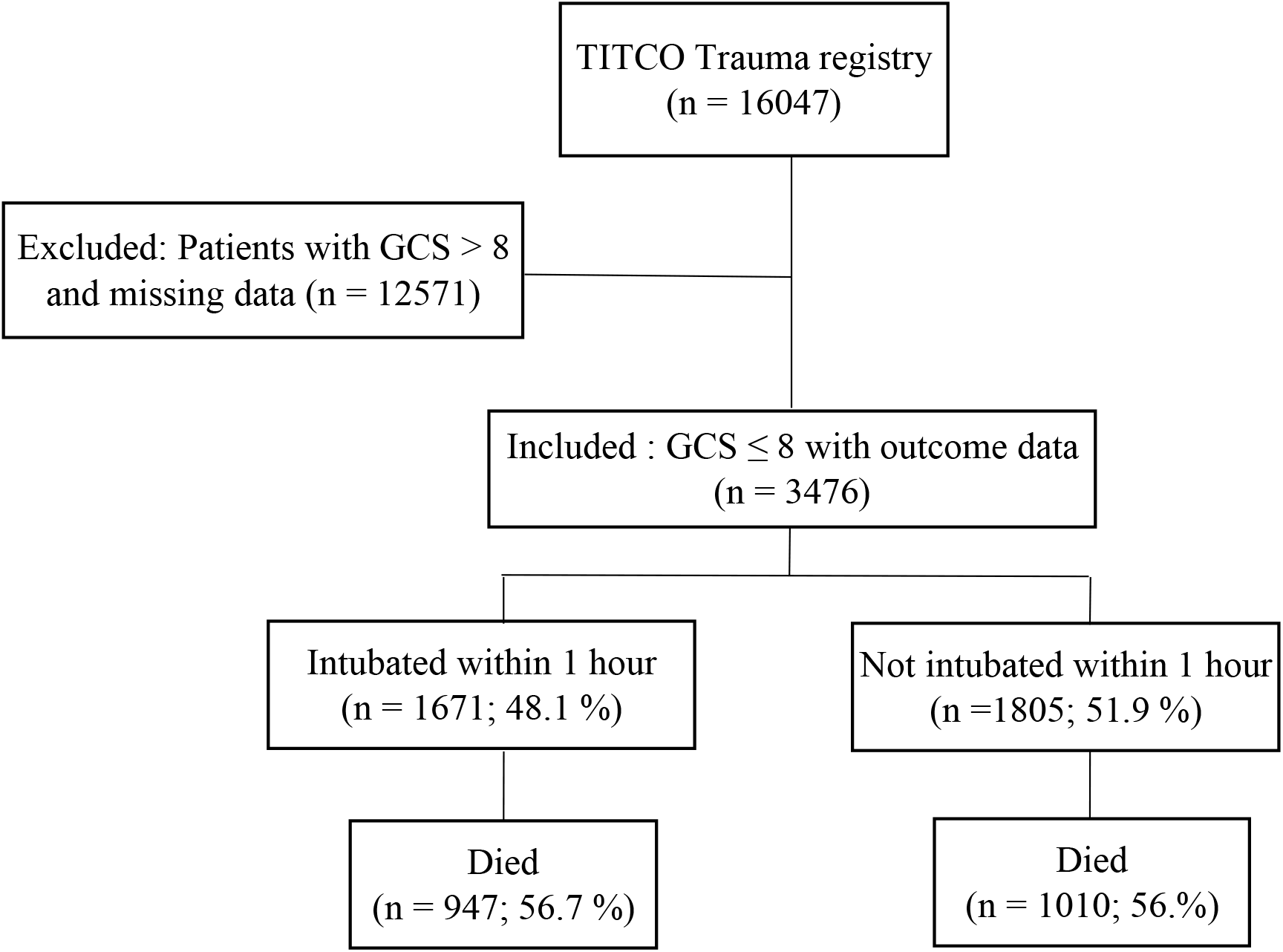
STROBE flow diagram for patient selection

Intubated patients were younger (30 [21.0 - 45.0] VS 35.0 [24.0 -52.0] years), more likely to be male (85.8 % VS 80.2 %), had higher ISS scores (14.0 [10 – 20.8] VS 10.0 [9.0 – 19.0]), and were more likely to arrive directly at the participating hospital rather than being transferred (transfer status 63.6 % VS 82.7 %) Road traffic injury was the most common mechanism in both groups (52.5 % in intubated and 57.5 % in non-intubated patients). Most injuries resulted from blunt mechanisms and ambulance was the most common mode of transfer in both groups. Intubated patients had higher HR (98.0 [86.0, 112.0] VS 90.0 [80.0, 100.0] beats/min), and lower RR (18.0 [16.0, 22.0] VS 20.00 [16.0, 24.0] breaths/min), SpO2 (96.8 [74.0, 100.0] VS 98.0 [96.5, 100.0] %), GCS score (5.0 [3.0 – 7.0] VS 6.0 [4.0 – 7.0],) and SBP at arrival (110.0 [96.8, 130.0] VS 110.0 [100.0, 128.0] mmHg) (Table 1).

**Table1 :** Baseline Characteristics of the patients who were intubated compared to those who were not in the Cohort

|  | Intubated within an hour | Not intubated within in hour | SMD |
| --- | --- | --- | --- |
| N | 1671 | 1805 |  |
| Age (median [IQR]) | 30.0 [21.0, 45.0] | 35.0 [24.0, 52.0] | 0.308 |
| Sex = Male (%) | 1433 (85.8) | 1447 (80.2) | 0.149 |
| Mechanism of injury |  |  |  |
| • Road traffic injury | 863 (52.5) | 1037 (57.5) | 0.359 |
| • Fall | 440 (26.8) | 536 (29.7) |  |
| • Railway injury | 252 (15.3) | 98 (5.4) |  |
| • Assault | 57 (3.5) | 63 (3.5) |  |
| • Burn | 1 (0.1) | 22 (1.2) |  |
| • Other | 31 (1.9) | 46 (2.6) |  |
| Penetrating trauma (%) | 25 (1.5) | 3 (0.2) | 0.147 |
| ISS (median [IQR]) | 14.00 [10.0, 20.8] | 10.00 [9.0, 19.0] | 0.165 |
| Transferred = Yes (%) | 1062 (63.6) | 1490 (82.7) | 0.443 |
| Mechanism of transfer (%) |  |  | 0.626 |
| • Ambulance | 984 (58.9) | 1487 (85.4) |  |
| • Car | 490 (29.3) | 156 (9.0) |  |
| • Taxi, motor rickshaw | 191 (11.4) | 94 (5.4) |  |
| • Other | 5 (0.3) | 5 (0.3) |  |
| SBP (median [IQR]) (mmHg) | 110.0 [96.8, 130.0] | 110.0 [100.0, 128.0] | 0.051 |
| SpO2 (median [IQR]) (%) | 96.8 [74.0, 100.0] | 98.0 [96.5, 100.0] | 0.678 |
| RR (median [IQR]) (/min) | 18.0 [16.0, 22.0] | 20.00 [16.0, 24.0] | 0.04 |
| HR (median [IQR]) (/min) | 98.0 [86.0, 112.0] | 90.0 [80.0, 100.0] | 0.242 |
| GCS (median [IQR]) | 5.0 [3.0, 7.0] | 6.0 [4.0, 7.0] | 0.286 |

In the analysis of cohort with complete outcome data, 947 (56%) patients out of 1671, had died in the intubation group whereas 1010 (56.7%) out of 1805 had died in the group of patients who were not intubated within an hour.

The patients who were intubated were ventilated for a median duration of 68.0 [10.0. 161.0] hours and had longer durations of ICU stay (78.0 [20.0, 173.0] VS 0.0 [0.0, 0.0] hours) and total hospital stay (6.0 [2.0, 15.0] VS 4.0 [1.0, 8.0] days) compared to those who were not. There was no significant difference between mortality in intubated (56.7 %) and non-intubated patients (56.0 %) (p = 0.695) on unadjusted analysis.

**Table 2:** Primary and Secondary Outcomes in whole cohort

|  |  |  |  |
| --- | --- | --- | --- |
| In-hospital mortality | 947 (56.7) | 1010 (56.0) | 0.695 |
| Length of ventilation (median [IQR]) (Hours) | 68.0 [10.0, 161.0] | 0.0 [0.0, 0.0] | <0.001 |
| Length of ICU stay (median [IQR]) (Hours) | 78.0 [20.0, 173.0] | 0.0 [0.0, 0.0] | <0.001 |
| Length of hospital stay (median [IQR]) (Days) | 6 [2.0, 15.0] | 4 [1.0,8.0] | <0.001 |

On multivariable logistic regression, greater age (OR = 1.03 [1.02, 1.04], p < 0.001) and ISS scores (OR = 1.04 [1.02, 1.07], p < 0.001), lower SBP (OR = 0.99 [0.98, 0.996]), p = 0.002) and GCS scores (OR = 0.57 [0.45, 0.71], p < 0.001) were significantly associated with in-hospital mortality whereas the intubation status (OR = 1.18 [0.76, 1.84], p = 0.467) was not.

**(Table 3).**

| Analysis: | Death | P-values |
| --- | --- | --- |
| Associations between Intubation and the End Point of Death in the Unadjusted Analysis, Multivariable Analysis, and Propensity-Score Analyses |  |  |
| Crude Analysis-odds ratio | 1.01 (0.96 - 1.07) | 0.695 |
| Multivariable Analysis | 1.18 (0.76-1.84) | 0.467 |
| Propensity Matching | 1.3 (0.73,2.36) | 0.403 |

The subgroup analysis based on GCS scores 3 to 5 and 6 to 8 revealed no significant difference (p value=0.421,p value= 0.133)in mortality in intubated patients compared to non-intubated patients. Though the overall mortality was higher in patients with GCS scores 3-5 (72.9 %) compared to those with scores 6-8 (39.5 %). Another subgroup analysis based on TBI and Non TBI also showed no significant mortality difference in intubated and non-intubated patients in both subgroups.Moreover, in the sensitivity analysis of direct patients with GCS ≤ 8, failed to show significant difference in mortality between both the groups (50.1% vs 49.9%, p value=0.11). The intubated patients had consistently longer durations of hospital stay and ICU admission across all subgroups., (SupplTables S5 and Figure 2).

A low GCS was most strongly associated with (OR = 0.78 [0.67-0.91], p = 0.002) the clinical decision to intubate, the others being a low SpO2 level (OR = 0.92 [0.89, 0.96], p < 0.001), high ISS (OR = 1.05 [1.01, 1.08], p = 0.012), and younger age (OR = 0.98 [0.971, 0.999], p = 0.03). SBP, RR, and HR were not significantly associated with intubation. (Table S6)

## Discussion

In this analysis involving a large cohort of trauma patients who had been hospitalized following acute trauma with Glasgow coma scale ≤ 8, the odds of inhospital mortality was not higher or lower among who received intubation within an hour than among those who did not (56.% vs 56.7%). In multiple analyses i.e unadjusted analysis, multivariable regression model (after adjusting for known prognostic factors) and on propensity matched control analysis, we found no significant association between intubation of patients arriving with GCS≤ 8 and inhospital mortality. We also performed a series of subgroup and sensitivity analysis using a priori hypothesis of heterogeneity of effect in different groups- GCS between 3 to 5, GCS between 6-8, patients with head injury, and without head injury.Sensitivity analysis was restricted to only directly admitted patients. However findings were similar across these analysis revealing no benefit for reduction of mortality.

The impact of intubation on survival in patients with severe GCS scores following trauma is largely disputed in recent studies. Hatchimoni and colleagues assessed 6676 adult patients with GCS 6–8 from the 2016 National Trauma Data Bank, 61.1 % of which were intubated within one hour of arrival (3). Intubation was independently associated with higher mortality in the overall cohort and subgroups with and without head injury. In a retrospective Trauma Quality Improvement Program study, including adult blunt trauma patients with GCS score of 7 or 8 and isolated head injury, Jakob et al. reported that 68.4 % patients were intubated immediately (within one hour of arrival)(17).In this study, immediate intubation was independently associated with 1.8 times higher odds of mortality whereas Andrusiek 6.8 times greater odds of developing pneumonia.(18)These findings were in line with study by Karamanos et al. in patients with iTBI and a GCS score of ≤ 8, in which pre-hospital endotracheal intubation was associated with a higher adjusted mortality rate, poor oxygenation, and a greater incidence of septic shock compared with oxygenation via face masks(8) These recent studies reporting greater mortality and morbidity amongst intubated patients may possibly reflect suboptimal patient selection for intubation and a large variations in intubation practices amongst centers (10,19) poor adherence to intubation protocols, and inadequate training of personnel (7). The safety of conservative management in patients with severe GCS scores, withholding intubation, has been demonstrated clearly only in certain cases of poisoning, but not in trauma settings. (20).On the contrary, The ProTECT III multicentre RCT showed prehospital intubation was associated with 47% lower odds of dying and better neurologic outcomes at six months compared with no intubation amongst patients with moderate to severe TBI, (21) Similarly Data from the pan-European cohort study, CENTER-TBI, demonstrated better outcomes with in-hospital intubation in patients with GCS scores ≤ 10 (19) These results suggest that the prevention of hypoxia and aspiration may outweighs the harms of intubation in selected TBI cases.

In our study we found that increasing age was independently associated with higher mortality in line with Jakob et al. (17), who reported 19.6 times greater odds of dying in patients aged ≥ 65 years, and Bick et al.(22) [Bick 2022], who reported more critical intracranial pathologies, trauma system activations, greater lengths of hospital and ICU stay, and mortality in patients aged ≥ 65 years. Hypotension, higher ISS scores, and low GCS scores were associated with higher mortality in our cohort, reflecting findings similar to previous studies (3,17,21). We also found that intubation was associated with longer length of hospital stay, ventilation, and ICU stay in the overall cohort and across all subgroups, in line with previous studies.(3,8,19,23).

We investigated the factors associated with intubation in our cohort, on exploratory regression analysis. The low GCS score was the strongest factor leading to the clinical decision of intubation. Others were younger age, low SpO2, and higher ISS scores. Earlier studies reported low GCS scores and younger age consistently to increasing the likelihood of intubation as well.Male sex, hypotension, and tachycardia were additional predictors of intubation(3,17,19). We considered only physiological variables in our model and deferred from including sex as it would be less likely to influence the decision to intubate as an independent factor in a clinical setting, irrespective of statistical differences.

### Strengths and limitations

The study reports the outcomes following intubation amongst trauma patients with severe GCS scores in a LMIC. It represent a large cohort from multiple tertiary care centers, representing samples from different geographic areas. Because the participating centers were fully equipped tertiary care centers that cater to severely injured trauma patients, they are representative of similar setups in most other LMICs. The multiple analysis and consistency of results across various subgroups, including sensitivity analysis, findings of the investigation do generate genuine uncertainty of overall benefit of intubation in the cohort of patients with GCS≤ 8.

Given the observational design with inherent limitations, the study should not be taken to rule out benefit or harm of intubation in trauma patients arriving with GCS ≤ 8.However, our findings do support the urgent need to re-evaluate recommendations from large trauma societies.

A randomized clinical trial is the best tool to determine efficacy of an intervention and whether benefit can be ascribed to the intervention under consideration. An observational trial has inherent issues of unmeasured confounding and bias. With the multiple analytical approaches we have attempted to limit the possible confounding in the investigation.

Despite these analytical adjustment, some unmeasured confounding may still persist. Details about how intubation was facilitated and the anesthetic drugs and sequence were not available in the registry. Additionally, the indication for intubation was not available in our dataset and may confound mortality statistics as some indications (eg. smoke inhalation, airway injury, risk of aspiration due to vomiting) are associated with higher mortality. A major limitation of the investigation is lack of information pertaining to inhospital complications, most notably development of chest infections and other sequalae which are often encountered in Trauma ICU such as ventilator associated pneumonia, and blood stream infections.

These complications are often difficult to capture in LMIC owing to inaccuracies in health records and reporting. Nevertheless, a prospective trial should consider them as important patient centered outcomes as these are associated with increased mortality and morbidity

Other possible confounding variables such as pre-existing comorbidities, intoxication, anticoagulation, and withdrawal of hospital care were either not collected or missing in the dataset in large quantities, and consequently were excluded from the analysis. Lastly, the data regarding long-term acute care admissions and long-term outcomes among patients requiring prolonged mechanical ventilation were not recorded in the registry because of limited resources for providing these facilities in the participating government hospitals (5).

## Conclusion

In our dataset, there was no significant difference in overall and subgroup (based on GCS-score stratification and injury location) in-hospital mortality in patients with severe GCS scores (3-8) intubated within one hour of arrival compared to those who were not intubated. Greater age and ISS, and lower SBP and GCS scores were independently associated with mortality in this group of patients. Intubation was associated with longer hospital stay, ventilation, and ICU stay. Low GCS scores, age, and oxygen saturation, and higher ISS scores increased the likelihood of the clinical decision to intubate.

## Data Availability

After signing a data-sharing agreement, the de-identified dataset is available upon request to the authors.

https://www.titco.org/

## Statements and Declarations

The authors have no competing interests. The study was approved by the Institutional Ethics Committee (IEC) of all participating hospitals - LTMGH (IEC/11/13 dated 26 Jul 2013), KEM (IEC (I)/out/222/14 dated 4 Mar 2014), SSKM (IEC/279 dated 21 Mar 2013, MMC (EC Reg no. ECR/270/Inst./TN/2013/36082013 dated 05.08.2014), and Apex Delhi (IEC/NP-327/2013 RP- 24/2013 dated 25 Sep 2013). The IECs individually approved the collation of the database and analysis, and granted waiver of individual consent for trauma patients, following which patients were enrolled with due written informed consent. The procedures used in this study adhere to the tenets of the Declaration of Helsinki.

## Availability of data and materials

After signing a data-sharing agreement, the de-identified dataset is available to researchers.

## Acknowledgment

We would like to acknowledge contribution of the participating hospitals and the staff in the TITCO research consortium of universities.

